# Assessing a multivariate model of brain-mediated genetic influences on disordered eating in the ABCD cohort

**DOI:** 10.1101/2022.10.02.22280578

**Authors:** Margaret L. Westwater, Travis T. Mallard, Varun Warrier, Richard A.I. Bethlehem, Dustin Scheinost, Christian Grillon, Paul C. Fletcher, Jakob Seidlitz, Monique Ernst

**Affiliations:** Department of Radiology and Biomedical Imaging, Yale University School of Medicine, New Haven, CT, USA; National Institute of Mental Health, National Institutes of Health, Bethesda, MD, USA; Psychiatric and Neurodevelopmental Genetics Unit, Massachusetts General Hospital, Boston, MA, USA; Department of Psychiatry, Harvard Medical School, Boston, MA, USA; Department of Psychiatry, University of Cambridge, Cambridge, UK; Department of Psychology, University of Cambridge, Cambridge, UK; Department of Psychiatry, University of Cambridge, Clifford Allbutt Building, Addenbrooke’s Hospital, Cambridge, UK; Wellcome Trust MRC Institute of Metabolic Science, University of Cambridge, Cambridge Biomedical Campus, Cambridge, UK; Department of Child and Adolescent Psychiatry and Behavioral Science, Children’s Hospital of Philadelphia, Philadelphia, PA, USA; Department of Psychiatry, University of Pennsylvania, Philadelphia, PA, USA; Lifespan Brain Institute of Children’s Hospital of Philadelphia and University of Pennsylvania, Philadelphia, PA, USA

## Abstract

Eating disorders (EDs) are complex psychiatric conditions that often emerge during adolescence, and affected individuals frequently demonstrate high rates of psychiatric comorbidity, particularly with depressive and anxiety disorders. Although risk for EDs reflects both genetic and neurobiological factors, knowledge of how genetic risk for EDs relates to neurobiology and psychiatric symptoms during critical developmental periods remains limited. We therefore implemented a novel multivariate framework, which sought to advance knowledge of the etiology of EDs by simultaneously estimating associations between genetic risk, brain structure and ED-related psychopathology symptoms in over 4,500 adolescents of European ancestry from the Adolescent Brain and Cognitive Development study (M(SD)_age_=119.29(7.49) months). Polygenic scores for anorexia nervosa (AN PGS) and body mass index (BMI PGS) were generated and related to three morphometric brain features— cortical thickness, surface area and subcortical grey matter volume—and to latent psychopathology factors using structural equation modeling. We identified a three-factor structure of ED-related psychopathology symptoms: eating, distress and fear factors. Increased BMI PGS were uniquely associated with greater eating factor scores, whereas AN PGS were unrelated to psychopathology factors. Moreover, genetic risk for high BMI and for AN had distinct neural correlates, where greater BMI PGS predicted widespread increases in cortical thickness and reductions in surface area while AN PGS were nominally related to reduced caudate volume. Altered default mode and visual network thickness was associated with greater eating factor scores, whereas distress and fear factor scores reflected a shared reduction in somatomotor network thickness. Our novel findings indicate that greater genetic risk for high BMI and altered cortical thickness of canonical brain networks underpin ED symptomatology in early adolescence. As neurobiological factors appear to shape disordered eating earlier in the life course than previously thought, these results underscore the need for early detection and intervention efforts for EDs.

## 1. Introduction

Eating disorders (EDs) are psychiatric conditions of complex etiology, in which individuals suffer with abnormal eating behavior, inappropriate weight loss behaviors (e.g., energy restriction, vomiting) and preoccupation with body shape or weight. These illnesses occur across the full weight spectrum, and they have a lifetime prevalence of 0.3—3.6% [1–3], where the majority of cases emerge during adolescence and follow a chronic, relapsing course [4–6]. While EDs are highly heritable, with genetic factors explaining 40-74% of phenotypic variation across anorexia nervosa (AN), bulimia nervosa (BN) and binge-eating disorder [7], knowledge of how genetic risk for EDs manifests across the lifespan remains limited. Genetic effects presumably influence one’s risk for psychiatric illness, in part, via their impact on the brain during critical developmental epochs, yet the specific relationships between genetic risk, neurodevelopment, and psychiatric outcomes have not been fully elucidated.

Emerging evidence from statistical genetics research suggests that the genes associated with AN, body mass index (BMI) and numerous psychiatric traits are primarily expressed in central nervous system tissue [8,9]. Additionally, genome-wide association studies (GWASs) of the human cerebral cortex have identified modest but significant genetic correlations between morphological features (e.g., surface area [SA], cortical thickness [CT], and grey matter volume [GMV]) and several neurocognitive traits, including neurodevelopmental disorders, major depressive disorder and depressive symptoms [10–12]. Related efforts have linked polygenic scores (PGS) for AN and BMI to altered SA in prefrontal and occipital regions, respectively, among adults [13,14]. This would suggest that genetic liability for mental illness and associated traits relates to morphometric features that can be readily estimated using non-invasive, *in vivo* neuroimaging. However, as these features change dynamically across the lifespan, attempts to integrate measures of genetic risk and brain structure must consider underlying developmental processes.

Adolescence represents a critical period that is characterized by not only rapid cortical maturation but also an increased incidence of psychiatric disorders [15–17]. Normative neurodevelopment follows a nonlinear trajectory, where average CT and GMV peak in early childhood at approximately 2 and 6 years, respectively, and decline in the second decade of life [18–20]. Total SA and subcortical volume continue to increase until adolescence, with the former peaking at age 11-12 and the latter ∼14 years. These alterations reflect a combination of synaptic pruning, axon elaboration and myelination that putatively facilitate further cognitive and social development during adolescence [21,22], and deviations from this trajectory have been reported in various psychiatric conditions [23,24], including AN [25,26] and BN [27].

Moreover, as genetic risk for psychopathology is developmentally dynamic, with new genetic influences coming ‘online’ across adolescence [28–30], the examination of gene-brain relationships during this period holds promise to advance understanding of the pathways by which genetic risk unfolds.

Such an approach has particular relevance to EDs, which often have a heterogeneous onset and extremely high rates of psychiatric comorbidity (71–94%), particularly with internalizing disorders like depressive or anxiety disorders [31,32]. Individuals suffering with EDs frequently demonstrate diagnostic crossover (e.g., from AN to BN), where most transitions occur within the first five years of illness [33,34]. Depressive symptoms, such as negative affect, often predate the emergence of disordered eating behaviors [35], and generalized anxiety during childhood has been shown to predict ED diagnoses in adolescence [36]. Moreover, premorbid obsessive-compulsive disorder (OCD) symptoms have been reported in AN [37], which may reflect their shared etiology as AN PGS has predicted OCD and anxiety symptoms in girls [38]. As such, efforts to improve risk stratification and early intervention for EDs would benefit from greater knowledge of the neural and behavioral correlates of elevated genetic risk for these illnesses and their related traits.

The present study sought to extend knowledge of the etiology of EDs by implementing a novel multivariate framework, which characterized associations among genetic factors, brain structure and ED-related symptoms in the Adolescent Brain and Cognitive Development (ABCD) study. Within this framework, we aimed to test whether PGS for AN and BMI were associated with ED, anxiety, depressive and obsessive-compulsive symptoms in youth; whether CT, SA and subcortical GMV explain variability in ED-related symptoms; and finally, whether the effects of AN and BMI PGS on ED-related symptoms are mediated by brain structure. These associations were examined in three multivariate models (i.e., one for each morphometric feature). We included the BMI PGS given that changes in body weight are a core diagnostic feature of several EDs (e.g., AN, atypical AN, avoidant and restrictive food intake disorder), and greater genetic risk for higher BMI has been related to disordered eating in other population-based cohorts [39,40]. By estimating gene-brain-behavior associations simultaneously, this approach enabled a more nuanced characterization of the shared and unique associations between ED-related symptoms and underlying neurobiology. We anticipated that BMI PGS would be associated with ED symptomatology, whereas the AN PGS would relate to ED and broader internalizing symptoms. We expected that ED symptoms would relate to distinct brain-behavior profiles as compared to other internalizing disorder symptoms, where the latter were predicted to have shared neural correlates. However, given the limited data on brain structure-psychopathology associations during early adolescence, we resisted making regionally-specific predictions. Finally, based on prior studies in adults [13,14], we anticipated that surface area would mediate the effects of AN and BMI PGS on psychopathology.

## 2. Methods

### 2.1 Participants

The initial sample included 11,867 children between the ages of 9 and 11 years (M(SD)_age_=9.91(0.62) years, n=5,681 female). Participants were recruited from one of 22 study sites, where recruitment efforts sought to enroll children of varying socioeconomic and ethnic backgrounds from all geographic census regions of the country [41]. Potential volunteers were primarily recruited through local schools at each site. Additionally, the ABCD Outreach and Dissemination Working Group used mailing lists, summer recruitment, twin registries and word-of-mouth referrals to reach interested individuals and their families.

### 2.2 Imaging Protocol

Imaging protocols for the ABCD cohort have been described previously [42], and acquisition parameters were harmonized across study sites and 3T scanner platforms (Siemens Prisma, General Electric 750 and Philips).

### 2.3 Cortical reconstruction and brain structure measures

Using FreeSurfer v6.0.1 software [43–45], cortical surface reconstructions were generated from raw T1-weighted anatomical images that were included in the curated ABCD Curated Annual Release 2.0 (doi:<u>10.15154/1503209</u>). Of the total sample, FreeSurfer reconstructions were available for 11,286 children. Morphometric estimates were extracted from the Desikan-Killiany parcellation [46] and seven bilateral subcortical regions (thalamus, caudate, putamen, pallidum, hippocampus, amygdala, nucleus accumbens). We then implemented a two-stage quality assurance procedure to exclude participants with low-quality surface reconstructions. First, we excluded children with neuroradiological findings of clinical significance, as well as those for whom significant image artifacts prevented radiological assessment (n=524). Second, we implemented an automated quality assurance step using FreeSurfer’s Euler number (i.e., a validated morphometric index of topographical complexity and technical image quality [47]), which was computed separately for each hemisphere and then averaged to yield a single value per subject. Participants (n=4) with Euler number values exceeding a previously validated cutoff and those with biologically implausible values (n=1) were excluded from subsequent analysis [47].

### 2.4 Genotyping procedures, imputation and quality control

Participants were genotyped using the custom Smokescreen™ genotype array, which contains over 300,000 single nucleotide polymorphisms (SNPs). Several stringent quality control steps were performed prior to imputation. First, SNPs with a genotyping rate <90% were removed. Then, individuals who had a genotyping rate <95%, whose genetic sex was discordant with their reported sex, and who had excessive heterozygosity were excluded. Hardy Weinberg Equilibrium and heterozygosity are not correctly calculated in ancestrally mixed populations, so we completed these steps in genetic ancestral groups that were identified using principal-component based clustering after participants’ data were combined with the 1000 Genomes Phase 3 data. Principal components of ancestry were estimated using GENESIS (http://www.imsbio.co.jp/RGM/R_rdfile?f=GENESIS/man/GENESIS-package.Rd&d=R_BC) after we accounted for relatedness between samples using KING software [48]. The first five principal components (PCs) were used to identify genetically homogenous groups in the 1000 Genomes data using UMAP, and this resulted in the identification of seven broad populations: Non-Finnish Europeans, Finnish Europeans, Africans, Americans, East Asians, South Asian, and Bengali. We then used the first five PCs from the ABCD dataset to project individuals onto these seven clusters, which identified largely homogeneous populations. Next, we implemented HWE based filtering (p<1×10^−6^) and excluded individuals with excessive heterozygosity (± 3 SD from the mean) as quality assurance steps. Genetic relatedness of participants in the final, cleaned dataset was estimated using KING software. PCs were re-calculated after accounting for relatedness, all at the level of individual population categories. We then merged, phased (Eagle v2.4) and imputed (Minimac4) these data using the TOPMED Imputation Server [49]. Finally, we removed SNPs with poor imputation (r^2^<0.4) and minor allele frequency <0.1% from the imputed data. Scripts for these quality control steps may be downloaded from: https://github.com/vwarrier/ABCD_geneticQC.

Since PGS have rather poor portability across ancestries [50], we generated PGS for AN and BMI in the 5,678 individuals of European ancestries in ABCD. Summary statistics were obtained from the largest prior GWAS of AN [8] and BMI [51]. We calculated PGS via Polygenic Risk Scoring using continuous shrinkage (PRS-CS), which has been shown to maximize the amount of variance explained by PGS computed from summary statistics relative to other methods [52]. This method also bypasses the need to identify a p-value threshold for SNP inclusion. Aligning with recommendations for highly polygenic traits, the global shrinkage prior (phi) was set to 1×10^2^. Resulting PGS for each participant were residualized for the first 10 PCs of ancestry, and they were Z-scored prior to analysis.

### 2.5 Regional and network-based measures of brain structure

We next computed regional and network-based estimates of cortical morphometry in the children of European ancestry, which served as explanatory variables in the subsequent structural equation models. CT and SA of the 68 cortical parcels and GMV of 14 subcortical regions were corrected for site effects using the neuroCombat (v1.0.13) and neuroCombatData (v0.99.1) R packages [53]. ComBAT models covaried for demographic, cognitive and developmental characteristics that have been previously associated with brain morphometry: age, sex [54,55], intelligence [56], years of parental education, pubertal development [57] and Euler number (see Supplementary Material). Euler number was included to further ensure that morphometric estimates were not influenced by the quality of the surface reconstruction [20]. Participants with missing data on any covariate (n=672) were removed via listwise deletion.

Individuals with extreme values (i.e., 5 SDs above or below the mean) across any adjusted ROI were classified as outliers and excluded from further analysis. To improve statistical power for neuroimaging inference [11,58] and structural equation modeling, adjusted regional CT and SA estimates were averaged and Z-scored across seven, functionally defined networks from the Yeo-Krienen atlas (**Figure 1a**; [59]). Adjusted subcortical GMV estimates were averaged across each hemisphere and Z-scored, yielding seven bilateral ROIs. The final samples included adjusted CT, SA and subcortical GMV measures from n=4,573, n=4,561 and n=4,547 children, respectively (see **Table 1**).

**Table 1.** Demographic and clinical information for final sample in each model

| Characteristic | Cortical Thickness Model (n = 4573) |  | Surface Area Model (n = 4561) |  | Subcortical GMV Model (n = 4547) |  |
| --- | --- | --- | --- | --- | --- | --- |
|  | M | SD | M | SD | M | SD |
| Sex (F/M) | 2128 / 2445 |  | 2125 / 2436 |  | 2124 / 2423 |  |
| Age (months) | 119.29 | 7.49 | 119.29 | 7.49 | 119.30 | 7.49 |
| BMI Z-score | 0.15 | 1.07 | 0.15 | 1.07 | 0.15 | 1.07 |
| Weight class<br>UW / HW / OW / OB | 215 / 3378 / 564 / 416 |  | 216 / 3370 / 560 / 415 |  | 216 / 3357 / 560 / 414 |  |
| Pubertal development | 1.50 | 0.41 | 1.50 | 0.40 | 1.50 | 1.40 |
| Fluid intelligence | 6.91 | 2.49 | 6.91 | 2.49 | 6.91 | 2.49 |
| Parental education (years) | 17.62 | 1.92 | 17.62 | 1.92 | 17.62 | 1.92 |
| Lifetime symptom endorsement | **N** | **%** | **N** | **%** | **N** | **%** |
| Eating disorders |  |  |  |  |  |  |
| *Emaciation* | 419 | 9.2 | 421 | 9.2 | 417 | 9.2 |
| *Fear of becoming obese* | 256 | 5.6 | 256 | 5.6 | 253 | 5.6 |
| *Objective binge-eating* | 172 | 3.8 | 171 | 3.7 | 171 | 3.8 |
| *Self-induced vomiting* | 6 | 0.1 | 6 | 0.1 | 6 | 0.1 |
| Depressive disorders |  |  |  |  |  |  |
| *Low mood* | 262 | 5.7 | 262 | 5.7 | 261 | 5.7 |
| *Anhedonia* | 207 | 4.5 | 208 | 4.6 | 208 | 4.6 |
| Anxiety disorders |  |  |  |  |  |  |
| *Difficulties controlling worry* | 644 | 14.1 | 645 | 14.1 | 641 | 14.1 |
| *Fear of social situations* | 532 | 11.6 | 531 | 11.6 | 528 | 11.6 |
| *Avoidance of phobic object* | 1623 | 35.5 | 1620 | 35.5 | 1617 | 35.6 |
| *Panic attack* | 228 | 5.0 | 227 | 5.0 | 225 | 4.9 |
| Obsessive-compulsive disorder |  |  |  |  |  |  |
| *Obsessions* | 610 | 13.3 | 607 | 13.3 | 606 | 13.3 |
| *Compulsions* | 385 | 8.4 | 387 | 8.5 | 386 | 8.5 |

**Note:** F = female, M = male, UW = underweight, HW = healthy weight, OW = overweight, OB = obese based age- and sex-adjusted BMI percentile.

**Figure 1.**
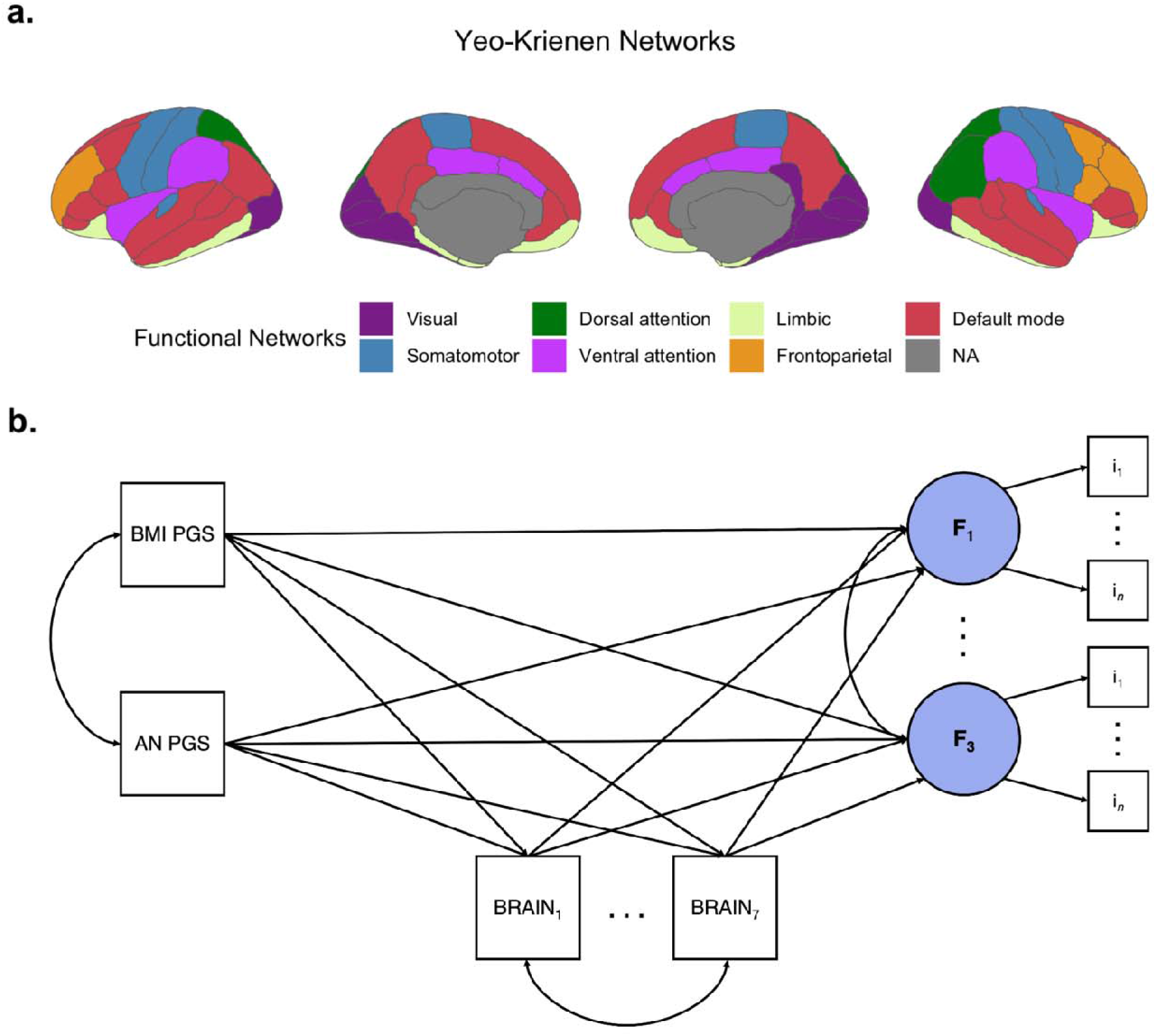
Simplified path diagram of multivariate model of gene-brain-behavior associations. Associations between AN and BMI PGS, brain morphometry and latent psychopathology factors were estimated using structural equation modeling. **a)** Cortical thickness and surface area estimates from the Desikan-Killiany parcellation were averaged across seven canonical Yeo-Krienen networks prior to inclusion in the SEM. **b)** Each model estimated the direct eff of explanatory variables (i.e., PGS and brain features) on latent psychopathology factors, which were identified from KSADS-5 item indicators. Moreover, the model estimated the cts indirect effects of PGS on latent psychopathology factors, as mediated by brain features. Note, each model included 7 brain features and 3 latent psychopathology factors with variable numbers of indicators; a simplified model was depicted here for illustration purposes. This model was estimated separately for each brain feature (cortical thickness, surface area, subcortical grey matter volume).

### 2.6 Behavioral measures and covariates

Children’s lifetime eating disorder, depressive, anxiety and OCD symptoms were derived from parent or guardian ratings on a validated, computerized clinical assessment (Kiddie Schedule for Affective Disorders and Schizophrenia; KSADS-5; [60]), which was optimized for the ABCD cohort (see [61]). Specifically, symptoms were assessed via screener items from eating disorder (emaciation, binge-eating, vomiting, fear of becoming obese), depression (low mood, anhedonia), anxiety (worry most days, fear of social situations, panic symptoms, active avoidance of phobic object) and OCD (obsessions, compulsions) modules. Details on symptom endorsement are provided in **Table 1** and **Figure S1**.

### 2.7 Structural equation modeling

Structural equation modeling (SEM) analyses were completed using MPlus software (v7; [62]). First, we estimated tetrachoric correlations among the 12 KSADS-5 items. Then, we performed confirmatory factor analyses, which tested three competing models: a common factor model and two correlated-factors models. The common factor model was selected on the basis of prior research, which suggests that ED, depressive, anxiety and OCD symptomatology may be explained by a higher-order ‘internalizing’ factor [63]. However, as others have found that ED pathology reflects a distinct subset of internalizing [64], we also tested a correlated-factors model, where one latent factor represented ED indicators, and a second factor represented depression, anxiety and OCD indicators. Finally, as depressive and generalized anxiety versus social anxiety, phobia, panic and OCD symptoms may reflect correlated but distinct ‘distress’ and ‘fear’ factors within the internalizing domain [64–66], we also tested a three-factor, correlated factors model that included latent ED, distress and fear factors. Item factor loadings were estimated using unit loading identification. Model fit was assessed using conventional indices generated by MPlus, including the model chi-square statistic, Comparative Fit Index (CFI), and root mean square error of approximation (RMSEA). Although a nonsignificant chi-square test indicates good model fit, this test is overpowered in large sample sizes, such as ABCD. CFI values above .90 suggest acceptable model fit, whereas RMSEA scores below .05 are indicative of good fit [67].

We then fit SEMs to estimate gene-brain-behavior associations across three morphometric features: bilateral subcortical GMV and average CT and SA across the Yeo-Krienen networks. This multivariate approach enabled the simultaneous estimation of associations between latent factors, their associated indicators, and the explanatory variables (i.e., PGS and brain morphometric measures; see **Figure 1b** for a simplified path diagram). Moreover, as regression estimates account for all other explanatory variables, these models facilitated the identification of specific gene-behavior, gene-brain, and gene-brain-behavior associations. All models were identified using weight least squares means and variance adjusted (WLSMV) algorithm with theta parameterization, which is suitable for categorical and nonmultivariate normal data. To account for the family relatedness of ABCD participants, individual observations were clustered within families (i.e., first-degree relatives) for all analyses. Alpha thresholds were adjusted for multiple comparisons across the three models using a Bonferroni correction (p = .05/3= 0.017), and results with corresponding p-values >0.017 but <0.05 were considered nominally statistically significant.

### 2.8 Sensitivity analyses

Our primary models included brain measures that were unadjusted for BMI as individual differences in brain structure have been related to both genetic and phenotypic indices of body mass [68–70], and genetic factors that dually influence BMI and the brain may be relevant to EDs. Therefore, we first estimated effects of BMI PGS on these measures without controlling for phenotypic effects of BMI. We then conducted sensitivity analyses, in which age- and sex-adjusted BMI Z-score was included as a covariate in our COMBAT models (see Section 2.5).

## 3. Results

### 3.1 Behavioral analyses support a three-factor structure of ED-related psychopathology

Tetrachoric correlation analyses indicated that KSADS-5 items were positively correlated with each other except for two items (lifetime emaciation and vomiting), which had weak or negative correlation estimates (**Figure S2 & Table S1**). As lifetime emaciation was largely uncorrelated with the other KSADS-5 items, it was excluded from subsequent analyses. Given the low endorsement of lifetime self-induced vomiting (n=6), this item was also excluded from downstream analyses. We then used confirmatory factor analysis to empirically test whether the observed covariance among remaining items could be explained by either a common or correlated-factors model. Fit indices for the common factor model suggested acceptable fit (CFIs=0.903-0.951, RMSEAs=0.047-0.05), but both the two- (CFIs=0.912-0.951, RMSEAs=0.046-0.048) and three-factor (CFIs=0.924-0.954, RMSEAs=0.046) solutions demonstrated improved model fit (see **Tables S2-S4** for fit information across CT, SA and subcortical GMV samples).

### 3.2 Cortical thickness

Fit indices from the CT model indicated that the model fit well and closely approximated the observed covariance matrix (χ^2^(df)=318.05(95), p<0.001, CFI=0.988, RMSEA=0.023). In this model, BMI PGS scores were positively associated with ED factor scores (B(SE)=0.314(0.035), p<0.001; **Figure 2 & Table S5**), and they were nominally related to greater CT across the ventral attention network (**Figure 3B**; B(SE)=0.032(0.015), p=0.031). However, average CT of the default mode (B(SE)=-0.288(0.104), p=0.006) and visual networks (B(SE)=0.177(0.062), p=0.004) were significantly associated with increased ED psychopathology scores. The directionality of this effect differed across the networks, where reduced default mode network thickness and increased visual network thickness conferred ED liability (**Figure 3B**). Moreover, lower CT of the somatomotor network was related to increased scores on both distress (B(SE)=-0.115(0.043), p=0.008) and fear (B(SE)=-0.146(0.042), p=0.001) factors. All effects of AN PGS on CT and psychopathology factors were nonsignificant.

**Figure 2.**
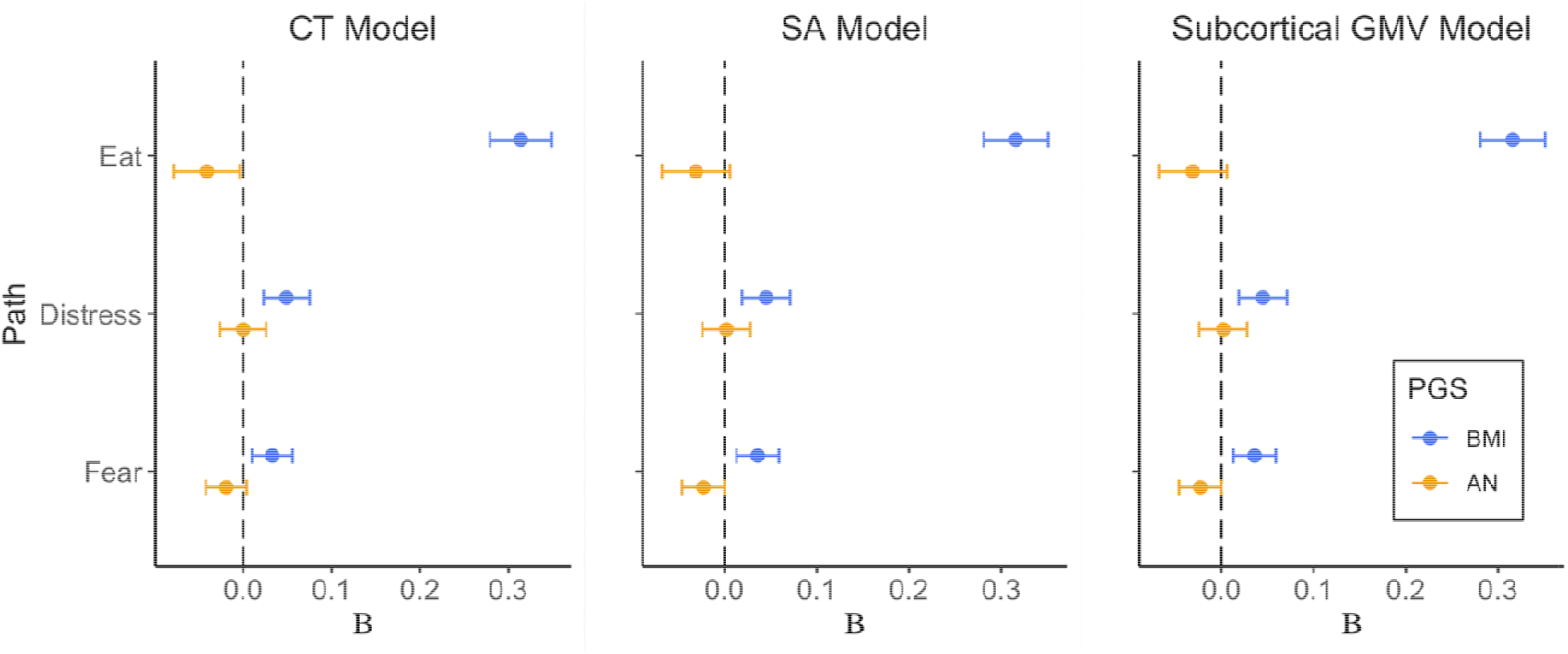
Genetic risk for high BMI is uniquely related to eating disorder psychopathology scores. Higher PGS for high BMI were associated with greater scores on the eating factor across all models (all p’s < .001). BMI PGS were nominally related to greater distress-related psychopathology in the Subcortical GMV model only (B(SE) = 0.054(0.023), p = 0.041). Values represent standardized beta estimates ± SEs from direct paths in the structural equation models. CT = cortical thickness, GMV = grey matter volume, SA = surface area.

**Figure 3.**
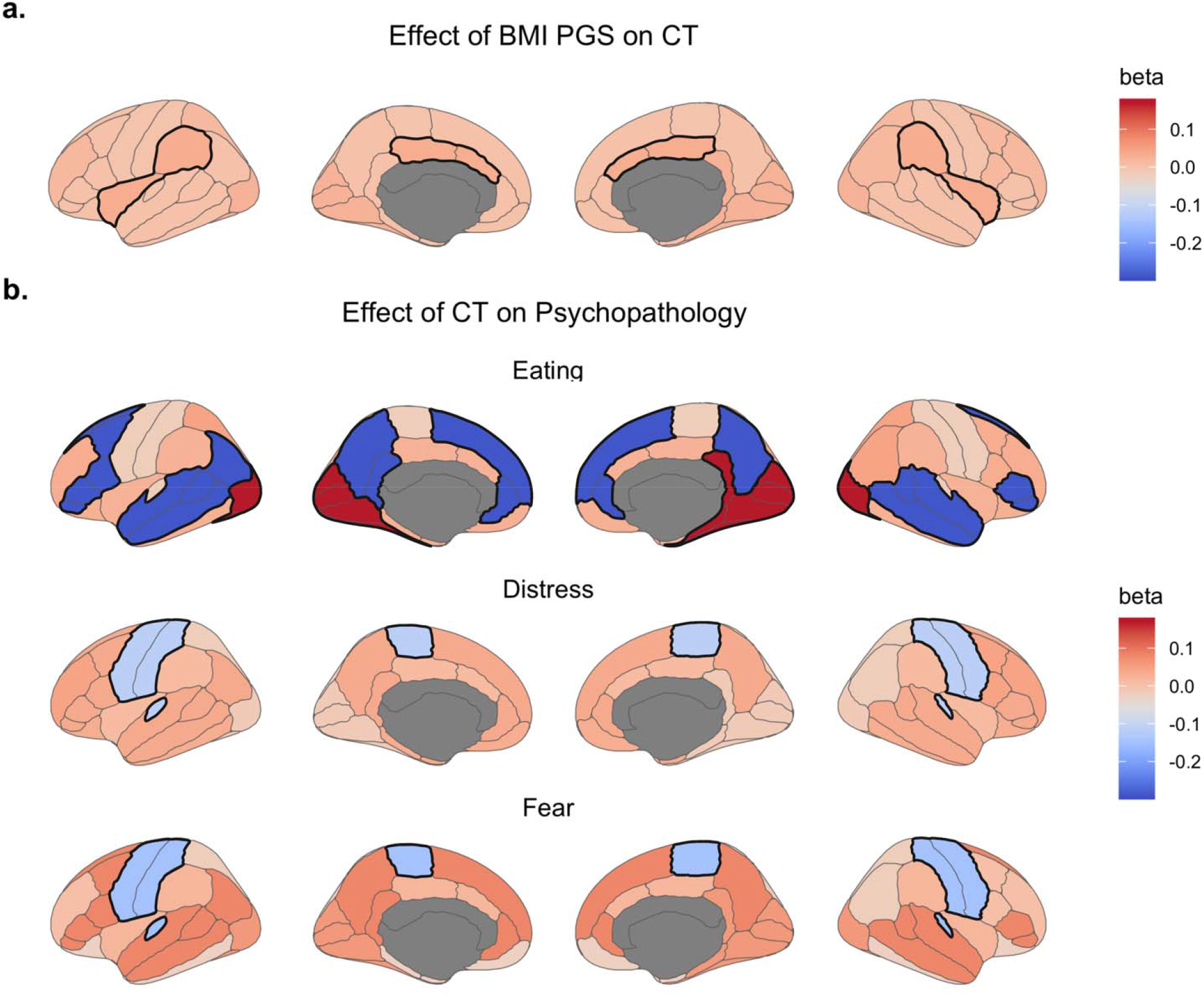
Associations between BMI PGS, cortical thickness and psychopathology. **a)** Effect estimates of associations between BMI PGS and cortical thickness. Increased BMI PGS scores related to greater average thickness of the ventral attention network. **b)** Effect estimates of associations between cortical thickness and latent psychopathology factor scores. Reductions in average thickness of the default mode network and increased thickness of the visual network were uniquely associated with eating psychopathology. However, lower average somatomotor network thickness was associated with both distress- and fear-related psychopathology. Statistically significant associations are denoted with a bolded border. BMI = body mass index; CT = cortical thickness; PGS = polygenic score.

### 3.3 Surface area

Fit indices from the SA model were also indicative of acceptable model fit (χ^2^ (df)=360.54(95), p<0.001, CFI=0.975, RMSEA=0.025). Aligning with the CT model, we observed a positive relationship between BMI PGS and ED psychopathology in the SA model (B(SE)=0.316(0.035), p<0.001; **Figure 2 & Table S6**). Moreover, increased BMI PGS scores were associated with widespread reductions in SA that were statistically significant in frontoparietal (B(SE)=-0.052(0.015), p=0.001), limbic (B(SE)=-0.043(0.016), p=0.005), and somatomotor (B(SE)=-0.042(0.016), p=0.007) networks and nominally significant in the ventral attention network (B(SE)=-0.031(0.015), p=0.042; **Figure 4A**). SA was unrelated to psychopathology factor scores, and AN PGS were not associated with SA or psychopathology factors.

**Figure 4.**
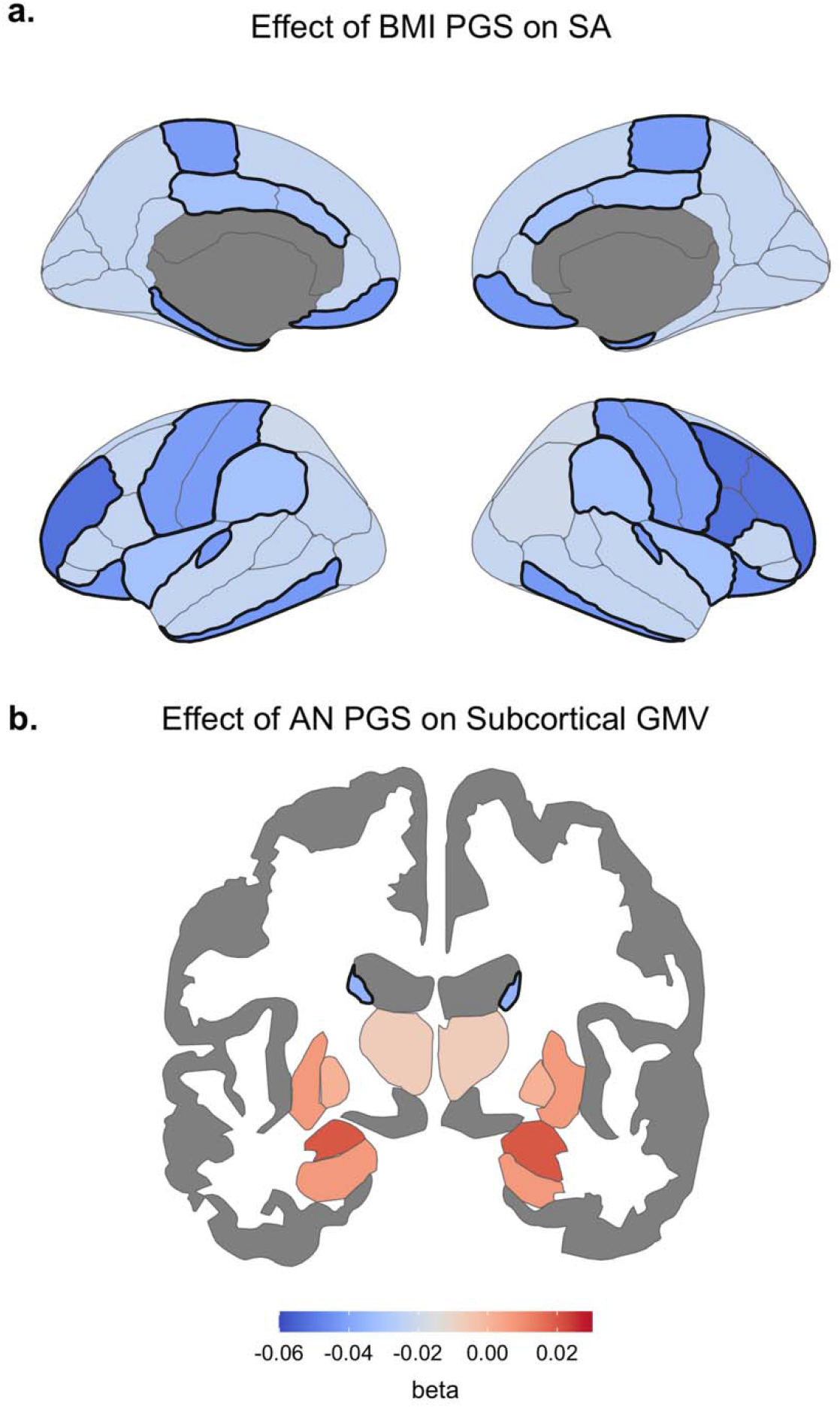
BMI PGS and AN PGS show unique associations with cortical morphometry. A) Standardized beta estimates of associations between BMI PGS and surface area across all Yeo-Krienen networks. Greater genetic risk for high BMI was associated with reduced surface area across ventral attention, limbic, frontoparietal and somatomotor networks. B) Standardized beta estimates from the Subcortical GMV model show that increased genetic risk for anorexia nervosa corresponded to lower bilateral caudate volume; however, this effect was nominally statistically significant (p=0.022). Significant networks or regions are depicted with a bolded border. AN = anorexia nervosa; GMV = grey matter volume; BMI = body mass index; PGS = polygenic score; SA = surface area.

### 3.4 Subcortical GMV

Similar to prior models, we found that the GMV model fit the data well (2χ (df)=396.34(95), p<.001, CFI=0.962, RMSEA=0.026). Greater genetic risk for high BMI was related to elevated ED (B(SE)=0.315(0.036), p=0.041) and distress factor scores (B(SE)=0.054(0.026), p=0.041), although the latter was nonsignificant following Bonferroni correction (**Figure 2 & Table S7**). AN PGS scores were nominally negatively associated with bilateral caudate volume (B(SE)=-0.036(0.016), p =0.022), whereas BMI PGS were not significantly related to subcortical GMV (**Figure 4B**). All associations between subcortical GMV and psychopathology factor scores were nonsignificant.

### 3.5 Sensitivity analyses

When CT, SA and subcortical GMV values were adjusted for BMI Z-score, we identified several distinct effects in SA and subcortical GMV models, whereas CT effects remained largely unchanged (see **Tables S8-S10**). In the BMI-adjusted SA model, BMI PGS scores were related to significantly lower SA in frontoparietal and limbic networks only, yet greater frontoparietal SA was nominally associated with increased ED factor scores (B(SE)=0.085(0.042), p=0.041). Moreover, BMI PGS were related to reduced bilateral thalamic volume (B(SE)=-0.039(0.023), p=0.016) when adjusting for BMI. Accounting for BMI Z-score did not affect associations between AN PGS and any brain or behavioral measures.

## 4. Discussion

In this study, we examined how genetic risk for AN and high BMI relates to individual differences in brain morphometry and eating disorder-related psychopathology in young adolescents of European ancestry. Using a theoretically informed SEM framework, we identified three key findings, which provide novel insight into the etiology of ED-related psychopathology during this critical period. First, genetic risk for high BMI was uniquely associated with ED psychopathology. Second, BMI PGS were related to alterations in average cortical thickness and surface area across several cortical networks, whereas genetic risk for AN was nominally related to reduced caudate volume. Finally, we provide novel evidence of associations between cortical thickness and ED psychopathology in early adolescence, which were distinct from the neural correlates of distress and fear psychopathology. We discuss the potential impact of these findings on adolescent mental health in the remaining sections.

### 4.1 Genetic risk for high BMI explains eating disorder psychopathology in young adolescents

We extend prior reports of positive associations between BMI PGS and disordered eating behaviors in adolescence [39,40], indicating, for the first time, that such associations are detectable in youth as young as 10 years of age. This parallels longitudinal observations at the phenotypic level, which indicate that adolescents with binge-eating/purging EDs have greater premorbid BMI relative to their unaffected peers [71]. Although BMI PGS were previously related to increased transdiagnostic psychopathology scores derived from the Child Behavioral Checklist in ABCD [72], we found limited support for an association with distress-related psychopathology, as measured by the KSADS-5. As associations between BMI PGS and other psychiatric traits have been primarily observed in older cohorts [73–75], it may be that such risk manifests across the second and third decades of life. Moreover, AN PGS were not associated with eating disorder-related psychopathology, and this may reflect various factors, including the developmental period under examination, the relatively low statistical power of the discovery AN GWAS compared to the BMI GWAS, or the specification of our ED factor, which was limited to two KSADS-5 items (lifetime binge-eating or fear of becoming obese) that do not capture core aspects of AN symptomatology. Nevertheless, our finding complements another report of nonsignificant associations between AN PGS and Child Behavioral Checklist scores in this cohort [76], which suggests that this null effect is not specific to the KSADS-5 items.

### 4.2 Polygenic scores for high BMI and AN are differentially associated with brain structure

As anticipated, both BMI PGS and AN PGS explained individual differences in brain morphometry. BMI PGS were related to greater average thickness of the ventral attention network, as well as reduced surface area of this network and much of the association cortex. Given that surface area peaks and cortical thickness begins to decline in early adolescence [20], our results may suggest that greater genetic risk for high BMI is associated with protracted brain maturation. This complements findings at the phenotypic level, linking increased BMI Z-score for age [77] and visceral fat mass [78] to greater cortical thickness, particularly of frontotemporal and occipital areas, in adolescence. Increased areal expansion and gyrification during development likely reflect a combination of cellular (e.g., synaptogenesis, gliogenesis, or intracortical myelination) and environmental (e.g., rearing environment) factors that facilitate increased functional integration of the cortex [79]. As such, regionally-specific negative associations between surface area and cortical thickness may arise from perturbations in these processes, particularly intracortical myelination [80]. Intracortical myelination is thought to underlie ‘stretching’ of the cortex tangentially along the surface [81], resulting in reduced thickness with increased surface area. Although knowledge of the effects of BMI-associated genetic loci on specific biological pathways is still developing, bioinformatic analyses have implicated these loci in synaptic functioning and neurotransmitter receptor activity [9], and modulation of these pathways could shape neurodevelopmental trajectories. Indeed, BMI PGS were previously related to lower occipital surface area in mid-adulthood [14], yet our findings indicate that this relationship may reflect altered neurodevelopment much earlier in life, which has implications for neurocognitive models of overweight and associated interventions.

However, as current PGS explain approximately 10% percent of phenotypic variation in BMI [51] which has itself been related to altered brain structure [68,69,77], it is important to consider that associations between BMI PGS and morphometric features may also covary with body mass. We therefore conducted sensitivity analyses, in which brain features were conditioned on BMI Z-score, and results largely recapitulated our primary models, albeit BMI

PGS were related to more localized reductions in surface area in frontoparietal and limbic networks only. This lends additional support to the notion that greater genetic risk for adiposity is related to delayed neurodevelopment, even when accounting for an adolescent’s BMI Z-score. We encourage future studies to investigate whether and how the relationship between BMI PGS and cortical morphometric features changes across the life course.

In contrast to the widespread associations between BMI PGS and morphometric features of the cortical mantle, AN PGS were nominally associated with reduced caudate volume only. Both adolescent and adult patients with AN demonstrate altered caudate activation during learning and decision-making tasks [82,83], and a recent meta-analysis from the ENIGMA AN Working Group reported widespread reductions in subcortical grey matter volume, including caudate volume, in acutely ill and partially weight-restored AN patients [84]. Although we studied a population-based cohort, our findings might suggest that the observed subcortical alterations in AN are partially attributable to genetic factors and may constitute a risk factor for this illness. Analysis of follow-up ABCD waves will be central to more precisely characterize the role of this PGS-caudate association in the development of EDs over time.

### 4.3 Distinct cortical thickness alterations underlie eating disorder and related psychopathology

Finally, our modeling approach provided novel insight into the shared and distinct neural correlates of ED-related psychopathology in the developing brain. Aligning with our expectations, fear- and distress-related psychopathology were associated with a shared reduction in cortical thickness of the somatomotor network. Greater depressive symptomatology has been associated with accelerated age-related thinning of frontal lobe regions, including the pre- and paracentral gyri, in youth aged 10 to 25 years [85]. However, our findings might implicate either premature thinning of these regions or restricted growth earlier in the life course in the development of distress- and fear-related psychopathology symptoms during adolescence.

Average thickness of the default mode and visual networks had opposing effects on ED psychopathology scores, where lower default mode and greater visual network thickness related to increased ED psychopathology. As normative cortical thinning in adolescence follows a caudal-rostral gradient [19,86], the directionality of these effects may indicate altered cortical maturation in the emergence of ED psychopathology. One explanation might be that later peak thickness of visual cortex, in combination with a premature peaking of default mode network thickness, increases an individual’s liability for disordered eating.

Altered functional connectivity of both networks and their constituent regions has been observed across the spectrum of EDs [87,88], yet findings of altered default mode network connectivity in AN remain more equivocal (reviewed by [89]). Moreover, cortical thickness reductions in women with variable BN symptom severity preferentially occur in areas with increased global structural connectivity, several of which comprise the default mode network. Adolescent girls and women with BN also have reduced thickness and volume of a key default mode network node, the posterior cingulate [90,91]. The default mode network shows unique age-related increases in structure-function coupling across adolescence, which are posited to reduce interference between this ‘task-negative’ network and task-positive networks that support executive functioning [92]. As such, deviations from this trajectory, in combination with altered visual network development, may impair one’s ability to suppress self-referential thoughts when responding to external stimuli, which could contribute to various ED symptoms (e.g., attentional difficulties due to concerns about food or altered perception of one’s body shape). Ongoing data collection efforts in ABCD, as well as younger cohorts like the Healthy Brain and Child Development Study, will be critical to characterizing these developmental processes and how they modulate risk for ED-related psychopathology.

Although we identified significant gene-behavior, gene-brain and brain-behavior associations, all indirect effects were nonsignificant, meaning that associations between BMI PGS or AN PGS and psychopathology factors were not mediated by brain structure. As noted above, these null effects could relate to various factors inherent to ABCD (e.g., developmental epoch, limited clinical phenotyping), and they warrant consideration of complementary neuroimaging measures (e.g., myelination or functional connectivity) in future studies.

### 4.4 Limitations and conclusions

Despite the strengths of our multivariate approach, several limitations should be acknowledged. First, our analyses were restricted to children of European ancestry, which limits the generalizability of our results. This was done as a precaution to attenuate the risk of spurious correlations and inaccurate conclusions, as polygenic scores currently have poor portability across populations (i.e., poor predictive performance when used in another ancestry than the original GWAS; [50,93]). Nevertheless, we underscore that determining the generalizability of these results across ancestral groups is an important next step for this work, and we support calls for greater inclusion of diverse samples in GWASs of eating disorders and related phenotypes. Second, our indices of ED, fear- and distress-related psychopathology were restricted to KSADS-5 screener items that assessed a limited number of symptoms. However, by modeling latent variables, we were able to aggregate information across individual items to generate more informative dimensional measures of psychopathology. Third, to increase statistical power for our complex multivariate model, we averaged cortical thickness and surface area estimates across seven brain networks, which resulted in lower spatial resolution. We justified this on the basis of prior work indicating that the genetic architecture of psychopathology has widespread effects across the cortex [10,94], as well as the advantages of the model over mass univariate approaches (i.e., simultaneous estimation of shared and unique neural correlates of ED-related psychopathology). Moreover, we averaged morphometric features across canonical functional networks that reflect the modular organization of the cortex for improved interpretability [58]. Finally, as puberty and gender identity have been associated with ED risk and illness trajectories, future study in follow-up ABCD releases should consider how such factors impact on the neurobiological mechanisms identified here.

In summary, we found that greater genetic risk for high BMI and altered cortical thickness of canonical brain networks is associated with ED symptomatology in early adolescence when accounting for commonly comorbid forms of psychopathology. This would suggest that neurobiological factors begin to shape individual differences in disordered eating by 10 years of age or earlier, which predates the reported age(s) of onset for EDs [4,95]. As such, our findings underscore the importance of improved screening and early intervention efforts for youth with disordered eating to ameliorate the long-term outcomes of these serious illnesses.

## Data Availability

This manuscript used publicly available data from the Adolescent Brain and Cognitive Development Study. All data resulting from the analyses are reported in the manuscript and supplementary tables.

## Supplementary information is available at MP’s website

### Author contributions

MLW, TTM, JS, and ME designed the study. VW computed polygenic scores and performed quality assurance checks of genetic data. RAIB and JS processed neuroimaging data. TTM estimated genetic relatedness of ABCD participants. MLW prepared neuroimaging and behavioral data and performed all structural equation modeling analysis, with assistance from TTM. DS, CG and PCF provided unpublished code and resources. MLW drafted the manuscript. All authors revised the manuscript and provided critical intellectual contributions.

## Acknowledgements

MLW was supported by the NIH-Oxford-Cambridge Scholars Program and NIH T32DA022975. TTM receives support from NIH T32HG010464. VW is supported by St. Catharine’s College Cambridge. R.A.I.B. received support from the Autism Research Trust. PCF receives support from the Bernard Wolfe Health Neuroscience Fund. JS was supported by NIH T32MH019112. Finally, CG, ME and MLW received funding from the Intramural Research Program at the NIMH (ZIAMH002798). Data were curated and analysed using a computational facility funded by an MRC research infrastructure award (MR/M009041/1) to the School of Clinical Medicine, University of Cambridge and supported by the mental health theme of the NIHR Cambridge Biomedical Research Centre. The views expressed are those of the authors and not necessarily those of the NIH, NHS, the NIHR or the Department of Health and Social Care. This work also utilized the computational resources of the NIH HPC Biowulf cluster (http://hpc.nih.gov).

We thank Ajay Nadig (Harvard/MIT MD-PhD Program, Harvard Medical School) for helpful discussions to support this work. We also wish to thank the ABCD participants, their families, and the researchers who collected these data. Data used in the preparation of this article were obtained from the Adolescent Brain Cognitive Development^SM^ (ABCD) Study (https://abcdstudy.org), held in the NIMH Data Archive (NDA). This is a multisite, longitudinal study designed to recruit more than 10,000 children age 9-10 and follow them over 10 years into early adulthood. The ABCD Study® is supported by the National Institutes of Health and additional federal partners under award numbers U01DA041048, U01DA050989, U01DA051016, U01DA041022, U01DA051018, U01DA051037, U01DA050987, U01DA041174, U01DA041106, U01DA041117, U01DA041028, U01DA041134, U01DA050988, U01DA051039, U01DA041156, U01DA041025, U01DA041120, U01DA051038, U01DA041148, U01DA041093, U01DA041089, U24DA041123, U24DA041147. A full list of supporters is available at https://abcdstudy.org/federal-partners.html. A listing of participating sites and a complete listing of the study investigators can be found at https://abcdstudy.org/consortium_members/. ABCD consortium investigators designed and implemented the study and/or provided data but did not necessarily participate in the analysis or writing of this report. This manuscript reflects the views of the authors and may not reflect the opinions or views of the NIH or ABCD consortium investigators. The ABCD data repository grows and changes over time. The ABCD data used in this report came from: <u>10.15154/1503209</u>. DOIs can be found at https://nda.nih.gov/abcd/abcd-annual-releases.html.

