## Supplementary material for "Assessing a multivariate model of brain-mediated genetic influences on disordered eating in the ABCD cohort": Figure S1

1. **Methods**
   1.1 *Covariate description*

Children completed the computerized Matrix Reasoning Task from the Weschler Intelligence Test for Children-V (WISC-V; [1]) within a larger executive function testing battery, and standardized scores were computed to approximate fluid intelligence. Pubertal development was indexed as the child’s mean score on the parent-rated Pubertal Development Scale (PDS; [2]). Although children also completed the self-report PDS, we elected to use the parent ratings as caregivers may have greater knowledge of overall developmental changes (e.g., growth spurt), including where the child falls in this process of change [3]. BMI was calculated from the average of three measurements of the child’s height and weight. Age- and sex-adjusted BMI Z-scores were computed using the R package ‘jBmi’ (<https://github.com/jbirstler/jBmi>) in reference to the Centers for Disease Control 2000 lookup tables. Participants with biologically implausible BMI Z-score values (n=19) were excluded.

2. **Results**
   2.1 *Cortical thickness sensitivity analysis*

When adjusting regional CT values for BMI Z-score, BMI PGS scores remained associated with ED psychopathology (B(SE) = 0.317(0.035), p < 0.001) and increased ventral attention network CT (B(SE) = 0.032(0.015), p = 0.036; see **Table S8**). Reductions in CT of the default mode network (B(SE) = -0.211(0.105), p = 0.044) and greater visual network CT (B(SE) = -0.145(0.062), p = 0.02) were associated with increased ED psychopathology scores. Lower CT of the somatomotor network remained associated with greater distress (B(SE) = -0.115(0.043), p = 0.008) and fear (B(SE) = -0.147(0.042), p = 0.001) factor scores. As in the primary model, fit indices suggested an acceptable fit of this model (χ^2^ (df) = 316.63(95), p < 0.001, CFI = 0.988, RMSEA = 0.023).

2.2 *Surface area sensitivity analysis*

When controlling for BMI Z-score, BMI PGS scores remained positively associated with ED factor scores (B(SE) = 0.317(0.035), p < 0.001), and they were negatively associated with SA across both limbic (B(SE) = -0.031(0.016), p = 0.047) and frontoparietal networks (B(SE) = -0.043(0.015), p = 0.006; **Table S9**). However, increased frontoparietal network SA was related to greater ED psychopathology scores (B(SE) = 0.085(0.042), p = 0.041). Fit indices suggested adequate model fit (χ^2^ (df) = 359.76(95), p< 0.001, CFI = 0.975, RMSEA = 0.025).

2.3 *Subcortical GMV sensitivity analysis*

Greater BMI PGS scores were related to greater ED (B(SE) = 0.316(0.036), p < 0.001) and distress (B(SE) = 0.053(0.026), p < 0.042) psychopathology scores (see **Table S10**). Moreover, increased genetic risk for higher BMI conferred reduced bilateral thalamic volume (B(SE) = -0.039(0.023), p = 0.016), and as in the original model, AN PGS scores were negatively associated with bilateral caudate volume at nominal statistical significance (B(SE) = -0.035(0.016), p = 0.023). This model demonstrated acceptable fit (χ^2^ (df) = 396.42(95), p < 0.001, CFI = 0.963, RMSEA = 0.026).

1. Wechsler D. Wechsler intelligence scale for children–Fifth edition (WISC-V): Technical and interpretive manual. Bloomington, MN: Pearson Clinical Assessment; 2014;

2. Petersen AC, Crockett L, Richards M, Boxer A. A self-report measure of pubertal status: Reliability, validity, and initial norms. J Youth Adolesc [Internet]. J Youth Adolesc; 1988 [cited 2020 Nov 2];17:117–33. Available from: http://www.ncbi.nlm.nih.gov/pubmed/24277579

3. Cheng TW, Magis-weinberg L, Williamson VG, Cecile D. A researcher’s guide to the measurement and modeling of puberty in the ABCD Study Ⓡ at baseline. PsyArXiv [Internet]. PsyArXiv; 2020 [cited 2020 Nov 2]; Available from: https://psyarxiv.com/4fv3k

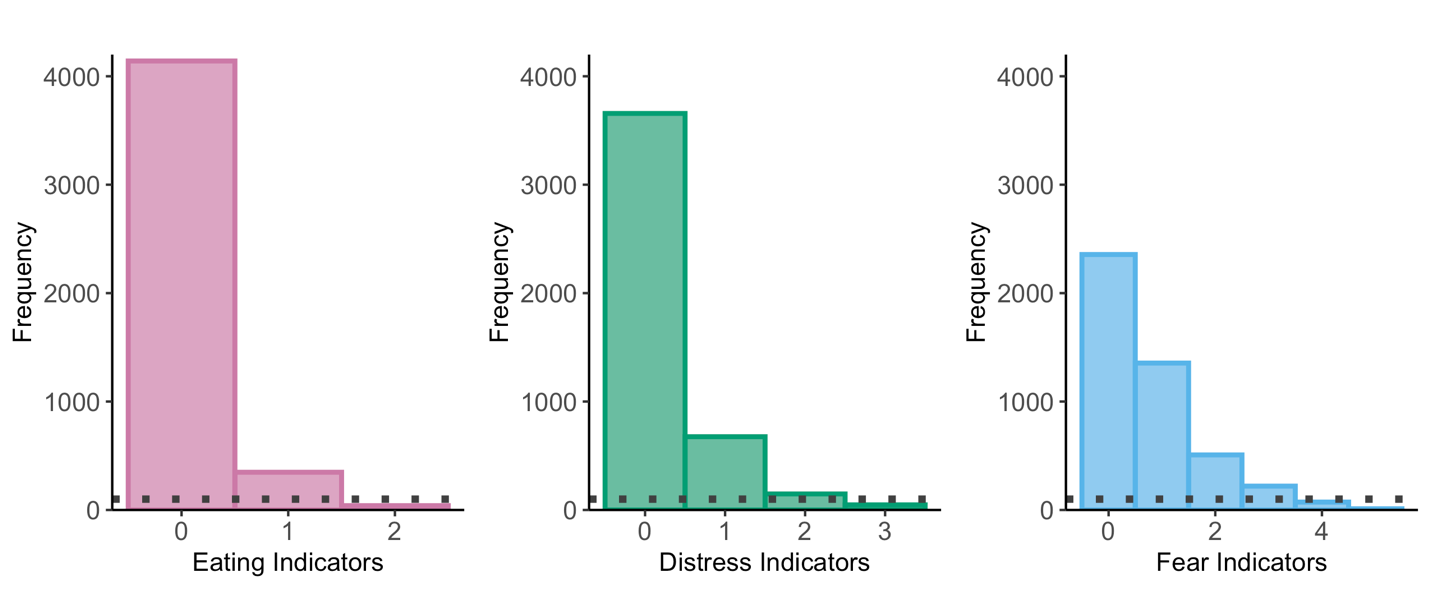

**Figure S1.** *Frequency of lifetime KSADS-5 item endorsement by factor.* To determine the number of eating disorder and internalizing psychopathology symptoms reported for each participant, item endorsement was summed for each latent factor. As would be expected, the distribution of sum scores for each factor was postively skewed, meaning that most parents reported one or fewer lifetime symptoms for their child. Histograms were generated in the full analytic sample for visualization. The dotted grey line indicates a frequency of 100.

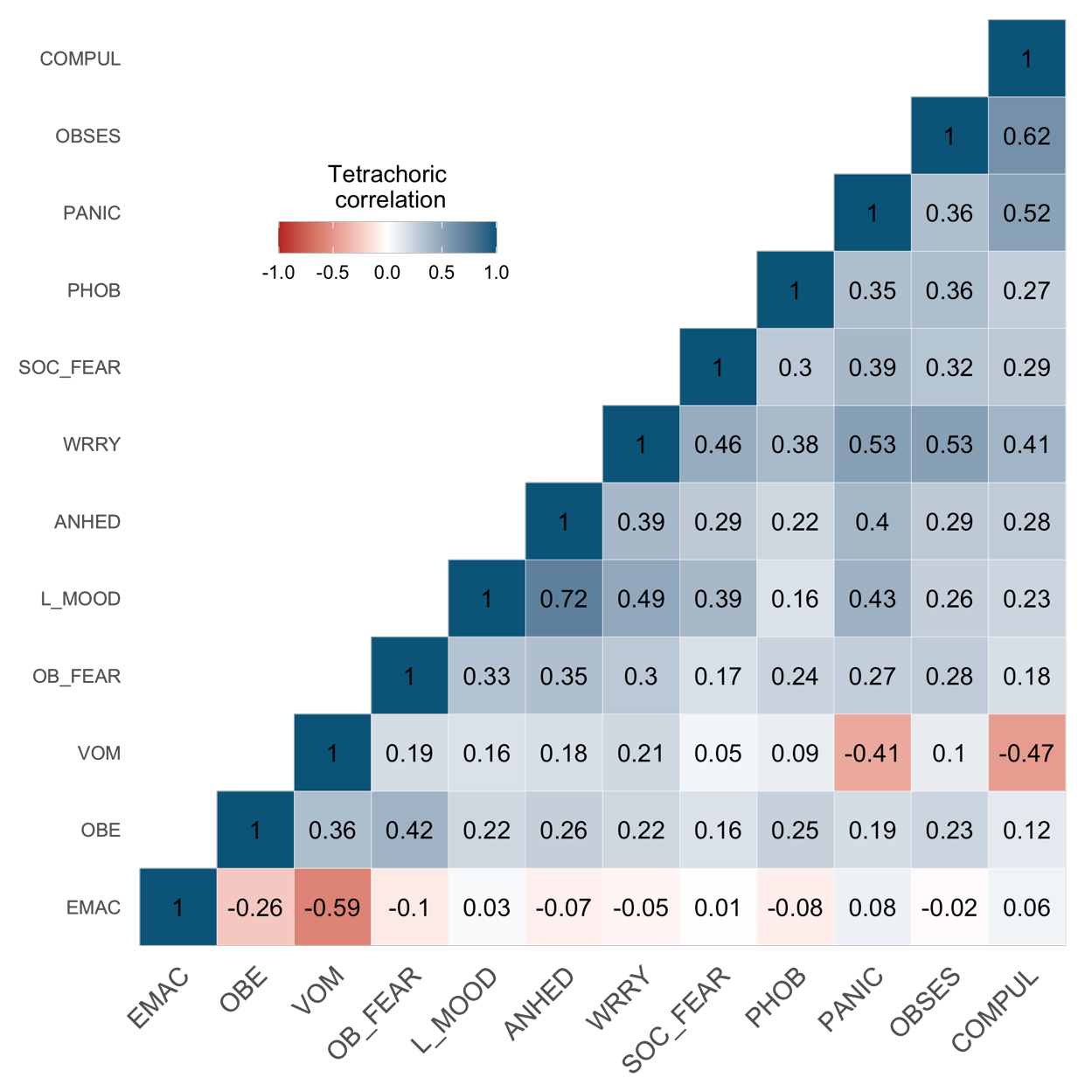

**Figure S2.** *KSADS-5 item correlations.* Items indexing lifetime internalizing and eating disorder psychopathology demonstrated moderate to strong postive correlations with one another; however, lifetime emaciation demonstrated weak, negative associations with most items (see Table S1 for corresponding p-values). Lifetime self-induced vomiting similarly was nonsignificantly correlated with other items. Correlation values were estimated in the maximum analytic sample for visualization. EMAC = emaciation, OBE = objective binge eating, VOM = self-induced vomiting, OB_FEAR = fear of becoming obese, L_MOOD = low mood, ANHED = anhedonia, WRRY = difficulty controlling worry, SOC_FEAR = fear of social situations, PHOB = avoidance of phobic object, OBSES = obsessions, COMP = compulsions
